# EEG signatures at different propofol vs sevoflurane concentrations

**DOI:** 10.1101/2024.01.24.24301740

**Authors:** C. Sun, A. Rigouzzo, I. Constant, D. Holcman

## Abstract

The depth of anesthesia is contingent upon the concentration of administered hypnotics, but establishing an exact relationship proves challenging, given its variability among individual patients. To elucidate the connection between the depth of anesthesia and hypnotic concentration, we leverage transient EEG patterns—specifically, iso-electric suppressions and power distributions within the *α* and *δ* frequency bands—at constant concentrations. Our investigation focuses on two hypnotic: propofol and sevoflurane. In a cohort encompassing children and young adults undergoing general anesthesia, we employ segmentation algorithms to extract a diverse range of spectral representations in EEG profiles. However, as we systematically alter hypnotic concentrations, a consistent trend emerges: heightened hypnotic concentration predominantly aligns with increased *δ*-band power and reduced *α*-band power. Notably, the occurrence of iso-electric suppressions is primarily associated with elevated propofol concentrations and infrequently observed with high levels of sevoflurane. Furthermore, we observe a decrease in the maximal power frequency of the *α*-band as hypnotic concentrations increase. In summary, this study offers a systematic quantification of EEG patterns corresponding to distinct concentrations of propofol and sevoflurane. These observed patterns contribute to a nuanced EEG representation of brain activity, laying the groundwork for personalized anesthesia strategies.

## 1 Introduction

Balance between patient comfort and clinical efficacy remains a challenge in modern anesthetic practices that requires a refined control of the hypnotic dose over time. Such control is achieved empirically by modulating the dose of classical hypnotic such as propofol and/or sevoflurane [1]. These prominent anesthesia agents are characterized by distinct molecular targets and mechanisms of action: while propofol is a GABAergic agonist, increasing the contribution of inhibition in the neuronal balance, sevoflurane is a NMDA antagonist [2], thus reducing excitation activity during synaptic neurotransmission. The choice between this intravenous agent propofol and volatile inhalation sevoflurane or both can introduce difficulties in monitoring the brain patient, due to their different EEG signatures [3, 1].

Non-invasive EEG recordings indicate brain electrical activity and allow monitoring the depth of anesthesia (DoA). The aim of this short study is to refine the relationship between electroencephalogram (EEG) signatures for various concentration steps of propofol and sevoflurane. EEG patterns include changes in *δ* waves during deep sedation to *α*and *β* oscillations in lighter sedation, a spectral representation that provide real-time insight into the cerebral response to anesthetic agents [4, 5]. Classifying these patterns obtained with propofol or sevoflurane at various concentrations would allow to obtain a better characterization of the heterogeneity brain responses across patients.

To investigate the relationship between EEG patterns at fixed propofol and sevoflurane concentrations in a cohort of children and young adults, we apply a spectral analysis to EEG recordings for various propofol and sevoflurane concentrations. We quantify below how the EEG frequency bands power are modified and the occurrence of specific patterns such as isoelectric suppressions or α*−*suppressions. This titration analysis can potentially be used to facilitate the interpretation of EEG responses and eventually be incorporated into automated and personalized anesthesia regimens based on individual EEG responses.

## 2 Materials and Methods

### 2.1 Statement about General Anesthesia protocol

This prospective observational randomized study included children and young adults patients receiving a scheduled elective procedure requiring general anesthesia under sevoflurane and propofol. We used a similar cohort as the one described in [6]. For each patient, two or more plateaus of 10 minutes duration were applied with a fixed concentration of hypnotic. This study was approved by the Ethics Committee CPP Saint Antoine, Paris, France (Approval number: 04605).

Written and informed consent was obtained from children and their parents. The protocol has been therefore performed in accordance with the ethical standards laid down in 1964 declaration of Helsinki and its later amendments. A group of 89 patients ranging from 6 to 38 years old, ASA 1 or 2, was used in this study. Data were collected between 2005 and 2006. The trial was later on registered and completed on September 2016 (ClinicalTrials.gov ID: NCT02893904). We used MATLAB R2021a software to perform the statistical analysis. The significance level used in this study was *α*=0.05. The values were expressed in mean *±* standard deviation.

### 2.2 General anesthesia protocol

All subjects received premedication with oral antihistamine hydroxyzine (1 mg.*kg*^*−*1^) before surgery. During induction, propofol was injected with an initial target concentration of 6 *μg*.*ml*^*−*1^. For sevoflurane, induction was performed at 6% with 100% oxygen. Both induction were followed by continuous administration of remifentanil (0.25 *μ*g.*kg*^*−*1^.*min*^*−*1^). The patients were then intubated with cuffed tracheal tube after a single dose curarization by atracurium besilate (0.5 mg.*kg*^*−*1^) and mechanically ventilated with an air-oxygen mixture and a breathing frequency ranging from 14 to 20 *min*^*−*1^ to obtain an *ET*_*CO*2_ between 30 and 35 mmHg. Propofol (resp. sevoflurane) concentration was kept between 2 and 6 *μg*.*ml*^*−*1^ according to the Schnider model for post-pubertal subjects and Kataria model for pre-pubertal subjects [7] (resp. 1 and 5%).

The decision to administer extra medication was left to the discretion of the anesthesiologist in charge following the institution standard of protocol care. The EEG monitoring was accompanied by Bispectral index (BIS), pulse oxygen saturation (SpO2), heart rate (CF), systolic and diastolic blood pressures (SBP, DBP), mean arterial pressure (MAP), and temperature estimation.

### 2.3 EEG recordings and pre-processing

EEG were recorded using the Brain-Quick program stem II (Micromed, France) with a single channel record from electrodes placed on the forehead, left and right (reference) mastoids [8]. We segmented the mechanical noise using the EEG signal power within a sliding window of length 10 seconds and 50% overlap. A time-segment *P*_*i*_ is considered artifactual when the spectral power within the window exceeds three times the median absolute value,

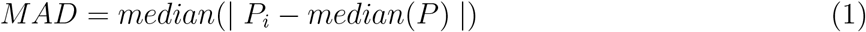

for *i* = 1, 2, .., *N*, where *N* is the number of windows. Segmented artifacts are then corrected using the classical Wavelet Quantile Normalization method [9, 10].

### 2.4 IES and *α*S segmentation

**Table 1:** Patients demographics, EEG and statistical features.

| Variables | 89 patients |
| --- | --- |
| Age (mean ± std) | 14.5 ± 7.2 yr |
| Age (range) | 6-38 yr |
| Gender (female/male) | 36 %/64 % |
| Pubertal (pre/post) | 56 %/44 % |
| Concentration plateau (propofol/sevoflurane) | 66/54 |

Iso-electric suppressions (IES) and *α−* suppressions (*α*S) were segmented following a procedure described in [4]: we first filtered the EEG signal *S*(*t*) in the range 8*−*16 Hz leading to the signal *S* _*α*_ (*t*). We normalized *S*_*α*_(*t*) by its Root-Mean-Square leading to *Ŝ* _*α*_ (*t*). We computed the difference *D*(*t*) and *D* _*α*_ (*t*) between the respective upper and lower envelops local maxima and minima interpolation, defining two threshold values

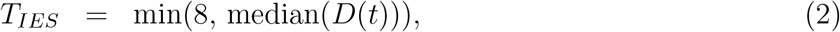

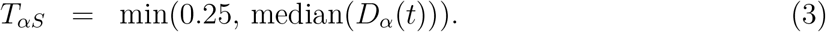

An EEG segment is classified as iso-electric (resp. *α*) suppression, when the amplitude of *Ŝ* _*α*_ is smaller than *T* _*αS*_, the amplitude of *S*(*t*) is smaller (resp. greater) than *T*_*IES*_ while the duration should exceed at least 1 second (resp. 0.5 second).

### 2.5 Mathematical indicators for the *α*and *δ* bands

The *α−* band relative power *Pα*(*t*) describes the power proportion of the concentration plateau in the [8, 12] Hz range with respect to [0.1, 45] Hz, as defined by the expression

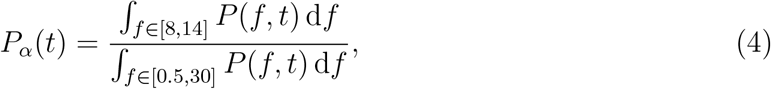

where *P* is the single channel EEG spectrogram obtained from a Fourier transform on a 40 seconds sliding window and 75% overlap. To obtain the mean value of the *α −*band relative power 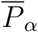 during the plateau duration, we apply the discretized sum approximation

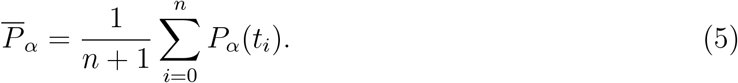

- The *δ−* band mean relative power 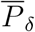 is obtained following the procedure above but taking [0.5, 4] Hz as the reference band.
- The maximum power frequency *f*_*α*_within the *α−* band is the frequency for which the power is maximal:

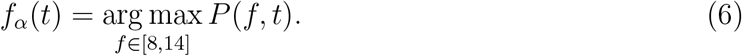

We then approximate the mean *α*_*max*_ by using the discretized sum

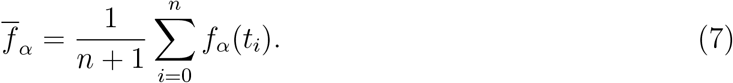

## 3 Results

### 3.1 EEG signatures at fixed propofol target concentration

We first presented (Fig.1A-B), EEG signatures and spectrograms for propofol and sevoflurane during two independent anesthesia. We further computed the proportion of suppressions of the alpha band (yellow) and iso-electric suppression (red) (Fig.1C-D), that can be compared with concentration of injected hypnotic (Fig.1E).

**Figure 1:**
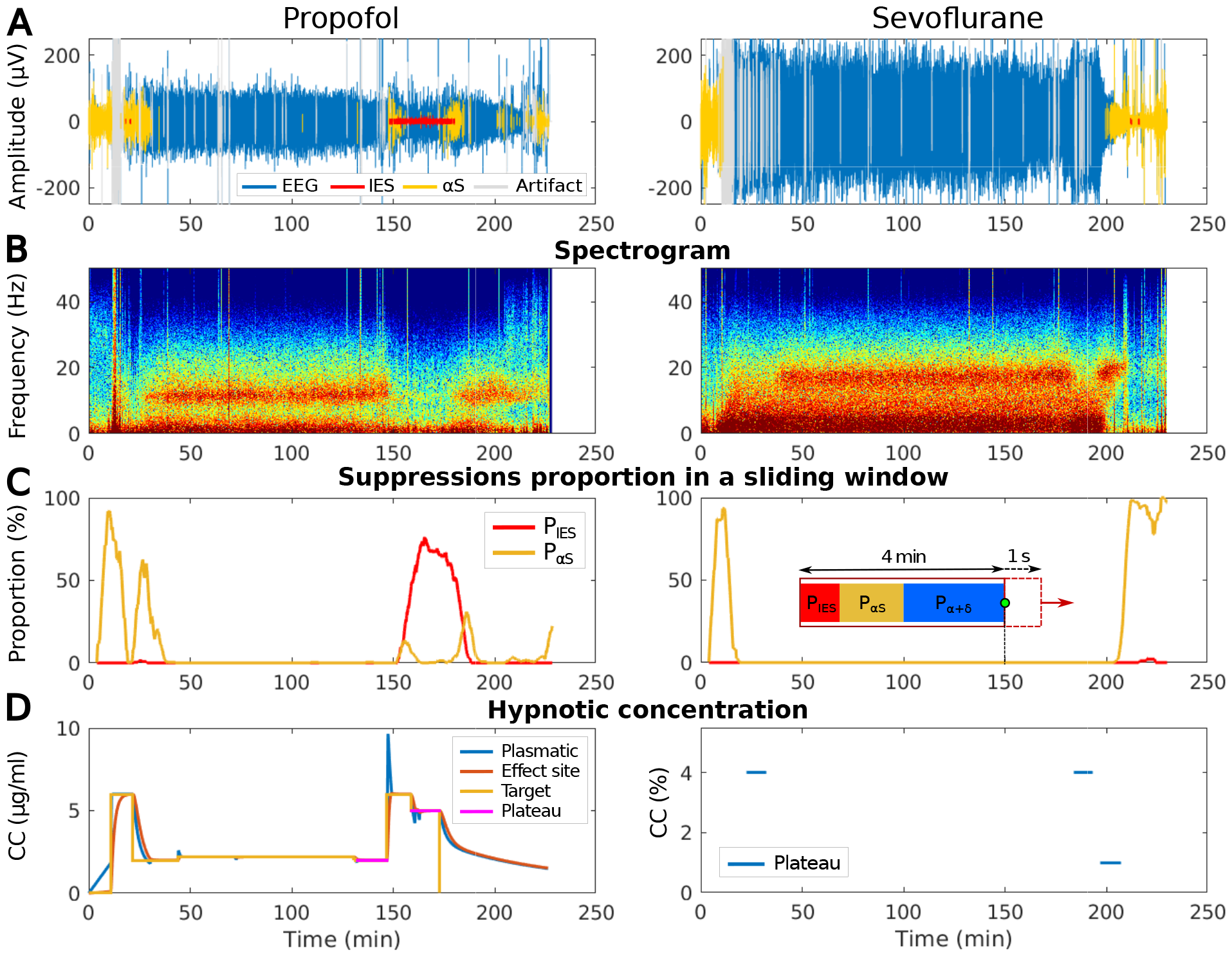
Difference of EEG waveform, spectrogram, suppressions proportion and hypnotic concentrations between propofol and sevoflurane induced general anesthesia. **(A)** EEG signal (blue) during propofol induced GA segmented in iso-electric suppressions (IES) (red), alpha-suppressions (*α*S) (yellow) and artifacts (grey). **(B)** Spectrogram (time-frequency) representation of the signal shown in (A). **(C)** Proportion of IES (red) and *α*S (yellow) evaluated in a 4 minutes sliding window and 1 second overlap. **(D)** Concentration of hypnotic injected. Propofol (left) has a target concentration (yellow) adjusted by hands, the plasmatic (blue), the effect site (orange) concentrations are computed according to pharmacokinetics and pharmacodynamics (PK/PD) models and the plateau (violet). For sevoflurane (right), only plateaus (blue) are shown.

To further characterize the EEG statistics, we computed the relative *δ−*band power *P*_*δ*_*/P*_*tot*_, where *P*_*tot*_ is the power computed across the frequency range 0.5-30 Hz. We found that the relative *δ −* band power increases along with the concentration (Fig.2A) for propofol, while it reaches a plateau for sevoflurane at an average of 3%. In parallel, the relative *α−* band power decreases as the concentration increases, reaching 5% of relative power when the concentration reaches 6 *μg*.*ml*^*−*1^ (Fig.2B). In addition, the relative *δ−* power also reached a concentration plateau at around 3% sevoflurane after decreasing. These results show a clear difference between sevoflurane vs propofol induced anesthesia.

**Figure 2:**
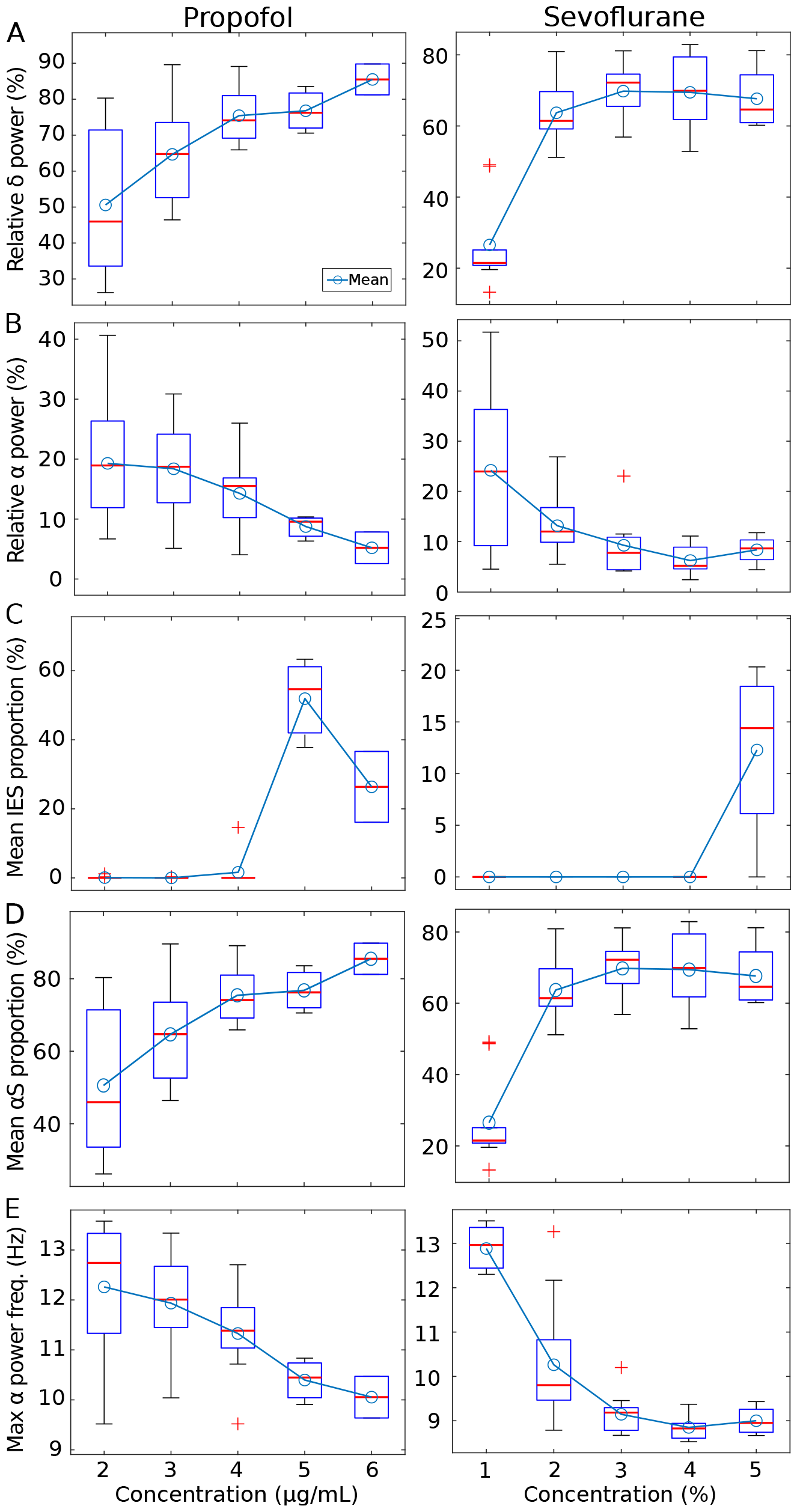
Distribution of IES, *α*S, band power versus propofol target concentration (left) and fraction of expired sevoflurane (right) plateaus. **(A)** Relative *δ−*band power distribution. **(B)** Relative *α−*band power distribution. **(C)** Mean IES proportion. **(D)** Mean proportion *α*S. **(E)** Average maximum power frequency within the *α−*rhythms range.

To further explore the arrival of deep sedation with concentration, we decided to detect the presence of iso-electric suppressions (IES) and suppression epochs of the alpha band (*α*S). We reported here a sudden increase of IES in both hypnotics. It is however not clear why the mean proportion of IES increases from 5 to 6 *μg*.*ml*^*−*1^ in the case of propofol. In parallel, we found that the mean proportion of *α*S increased continuously with respect to propofol concentration in the [2-6] *μg*.*ml*^*−*1^ range. In the case of propofol, *α*S increased to a maximum after 3% of sevoflurane. At this stage, there was no clear marks of anticipating IES from the dynamics of *αS* (Fig.2C). This increase is not necessarily associated with an increase in the mean IES proportion. Instead, IES appeared predominantly for concentration starting at 5 to 6 *μg*.*ml*^*−*1^. Finally, we observed that the mean of the frequency associated to the maximum power of the *α*band decreased continuously from 12.3 to 10 Hz (Fig.2E) as propofol concentration increases from 2 to 6 *μg/ml*, a situation that is comparable with sevoflurane.

### 3.2 EEG spectral signatures at fixed fraction of hypnotic

To explore the diversity of the spectral signatures of propofol vs Sevoflurane, we decided to plot the spectrograms at fixed fraction of hypnotic concentration (Fig.3): While for low concentrations, the spectrogram profile can be different across patients, we found that the SEF95 and median frequency was statistical different for the two categories of hypnotic. Indeed, the median frequency is in average higher for sevoflurane ([13.9, 3.0, 2.8, 2.9, 2.8] Hz) from 1 to 5%) compared to propofol ([7.0, 2.8, 1.4, 1.1, 1.0] Hz from 2 to 6 *μg*.*ml*^*−*1^) for all concentrations. For concentrations at 3 and 4 units, the SEF95 are in average lower for sevoflurane ([10.9, 9.6] Hz) against propofol ([12.3, 13.9] Hz).

**Figure 3:**
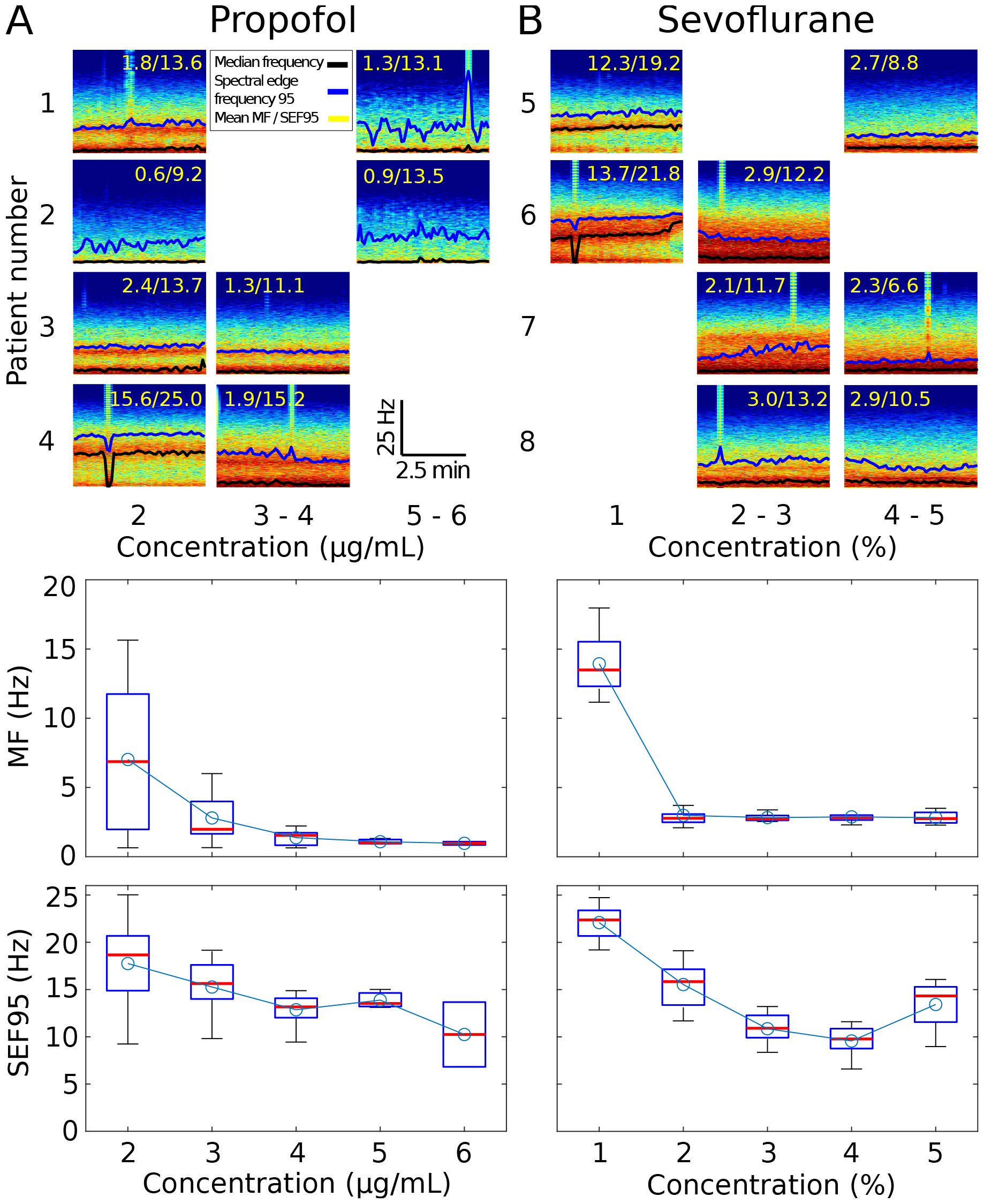
Spectrogram profiles with Median Frequencies (MF) and Spectral Edge Frequency 95 (SEF95) at different plateaus concentration for. **(A)** Propofol, **(B)** Sevoflurane. The Median Frequency (black) and spectral edge frequency 95 (blue) are shown over the spectrogram along with their mean values (yellow). The corresponding boxplots are shown below.

## 4 Discussion and conclusion

In this manuscript, we highlighted the differences between propofol and sevoflurane anesthetic concentrations and presented also the spectral properties of the EEG signal. We computed several parameters, such as the power of the *α*and *δ* bands for various sevoflurane and propofol concentrations. To evaluate the depth of anesthesia, we also computed the fraction of time in *α−*suppressions and isoelectric suppressions (Fig.2A-B). When propofol concentration increased, the power of the *α−*band decreased, while the *δ* power becomes dominant (Fig.2C-D).

However, we did not detect any general trends where the emergence of *α*S would systematically anticipate the appearance of IES. It seems that IES suddenly appear for a specific concentration, suggesting that at a hypnotic concentration high enough, a switch could occur of the dominant brain oscillations. Finally, the spectrograms further reveal a clear variability across patients (Fig.3).

When we compared propofol vs sevoflurane effect on EEG, we found no particular signature differences at low concentration of both hypnotic with a high variability between the spectrogram profiles. At medium concentration ([3-5]*μg*.*ml*^*−*1^ vs. [2-4]%), the higher median frequency (MF) but lower SEF95 for sevoflurane is associated with wider *δ* and *α*bands compared to propofol, where we have a lower MF but higher SEF95 that we associated with narrow bands and thus less signal amplitude at the corresponding bands frequencies. For high concentration, the higher MF and SEF95 for sevoflurane is associated with a more dominant *δ−*band, while for propofol the signal amplitude is more suppressed at *δ* frequencies. Finally, the present study complement previous studies [6, 11] on propofol and sevoflurane comparison.

## Abbreviations

EEG: ElectroEncephaloGram
GA: General Anesthesia
TCI: Target Controlled Infusion
*α*S: Alpha-Suppression
IES: Iso-Electric Suppression;

## Acknowledgments

D.H. research is supported by “Fondation pour la Recherche Médicale” (Postdoctorat en France - SPF201909009284), an ANR grant NEUC-0001 and the European Research Council (ERC) under the European Union ‘s Horizon 2020 research and innovation programme (grant agreement No 882673) and a CNRS Pre-maturation Grant.

## Data availability

The datasets generated and analysis in the current study are available from the corresponding authors upon reasonable request.

## References

[1] P. L. Purdon, A. Sampson, K. J. Pavone, and E. N. Brown, “Clinical electroencephalography for anesthesiologists: part i: background and basic signatures,” Anesthesiology, vol. 123, no. 4, pp. 937–960, 2015.

[2] M. T. Alkire, A. G. Hudetz, and G. Tononi, “Consciousness and anesthesia,” Science, vol. 322, no. 5903, pp. 876–880, 2008.

[3] I. Constant and N. Sabourdin, “The eeg signal: a window on the cortical brain activity, “Pediatric Anesthesia, vol. 22, no. 6, pp. 539–552, 2012.

[4] C. Sun and D. Holcman, “Combining transient statistical markers from the eeg signal to predict brain sensitivity to general anesthesia,” Biomedical Signal Processing and Control, vol. 77, p. 103713, 2022.

[5] P. L. Purdon, E. T. Pierce, E. A. Mukamel, M. J. Prerau, J. L. Walsh, K. F. K. Wong, A. F. Salazar-Gomez, P. G. Harrell, A. L. Sampson, A. Cimenser, et al., “Electroencephalogram signatures of loss and recovery of consciousness from propofol,” Proceedings of the National Academy of Sciences, vol. 110, no. 12, pp. E1142–E1151, 2013.

[6] A. Rigouzzo, L. Khoy-Ear, D. Laude, N. Louvet, M.-L. Moutard, N. Sabourdin, and Constant, “Eeg profiles during general anesthesia in children: A comparative study between sevoflurane and propofol,” Pediatric Anesthesia, vol. 29, no. 3, pp. 250–257, 2019.

[7] A. R. Absalom and K. P. Mason, Total intravenous anesthesia and target controlled infusions. Springer, 2017.

[8] A. Rigouzzo, L. Girault, N. Louvet, F. Servin, T. De-Smet, V. Piat, R. Seeman, I. Murat, and I. Constant, “The relationship between bispectral index and propofol during target-controlled infusion anesthesia: a comparative study between children and young adults,” Anesthesia & Analgesia, vol. 106, no. 4, pp. 1109–1116, 2008.

[9] M. Dora and D. Holcman, “Adaptive single-channel eeg artifact removal for real-time clinical monitoring,” IEEE Transactions on Neural Systems and Rehabilitation EngineeringF, 2022.

[10] M. Dora, S. Jaffard, and D. Holcman, “The wqn algorithm to adaptively correct artifacts in the eeg signal,” Applied and Computational Harmonic Analysis, vol. 61, pp. 347–356, 2022.

[11] O. Akeju, M. B. Westover, K. J. Pavone, A. L. Sampson, K. E. Hartnack, E. N. Brown, and P. L. Purdon, “Effects of sevoflurane and propofol on frontal electroencephalogram power and coherence,” Anesthesiology, vol. 121, no. 5, pp. 990–998, 2014.

